# Reliability-Aware Clinical Relation Learning for Uncertainty-Robust Multi-Label Chest X-ray Classification

**DOI:** 10.64898/2026.11.11.26367867

**Authors:** Yifan Jin, Tao Liu, Mark Stephen

## Abstract

**Background:** Automated multi-label classification of chest radiographs is complicated by overlapping imaging findings, noisy report-derived labels, and changes in disease co-occurrence across clinical settings. Existing clinical-hierarchy methods can encode label relationships, but a rigid hierarchy may conflate categorical membership with empirical association and propagate annotation errors.

**Methods:** We investigated Reliability-Aware Clinical Relation Learning (ReCaRL), a graph-assisted framework for multi-label chest radiograph classification. An image encoder produces initial finding scores, while a graph constructed from training-label associations provides reliability-weighted, confidence-gated updates. Uncertain annotations are handled with observation masks. The evaluation compares flat, weighted, graph-based, and hierarchy-penalty baselines, with internal and external discrimination and calibration assessments.

**Results:** In the internal evaluation, ReCaRL achieved macro AUROC 0.821 and macro AUPRC 0.437, compared with 0.812 and 0.423 for the masked/weighted baseline. Mean AUROC across five common findings was 0.865 versus 0.858. External testing yielded AUROC 0.799 and AUPRC 0.409. Post-hoc calibration reduced the internal Brier score from 0.113 to 0.102 and the external expected calibration error from 0.082 to 0.061, without changing AUROC or AUPRC.

**Conclusions:** Reliability-aware graph propagation and confidence gating were associated with modest improvements in multi-label chest radiograph classification relative to the matched masked/weighted baseline. Post-hoc calibration improved probability reliability in the reported evaluations. The findings do not establish a benefit in prospective clinical decision-making.

## 1 Introduction

Chest radiography is a high-volume examination used to assess diverse cardiopulmonary findings, including atelectasis, consolidation, edema, cardiomegaly, and pleural effusion. Contemporary public datasets have enabled the development of multi-label image classifiers at scale. CheXpert contains 224,316 chest radiographs from 65,240 patients and provides report-derived labels that distinguish positive, negative, and uncertain observations [1]. MIMIC-CXR includes 377,110 radiographs across 227,835 studies with accompanying radiology reports and provides an important independent institutional setting for model assessment [2].

Multi-label CXR prediction differs from a collection of unrelated binary tasks because findings may coexist and may be linked by anatomical or semantic structure. However, not all clinical relationships are equivalent. Categorical membership can be definitional (for example, pleural effusion belongs to a pleural-finding group), whereas associations among edema, effusion, and cardiomegaly are population-dependent and do not imply deterministic disease relationships. Directly converting such associations into hard parent-child constraints risks creating inconsistent targets or amplifying spurious correlations in unfamiliar patient populations.

A recent clinically inspired hierarchical approach combined DenseNet121 with a penalty-based hierarchical binary cross-entropy objective [3]. Earlier work also formulated conditional and unconditional predictions within chest radiograph taxonomies and examined learning from incomplete annotations [15]. Its reported overall hierarchy-level mean AUROC was approximately 0.903; notably, the original paper also reported a five-label flat model that performed better than several hierarchy-penalty configurations [3]. This distinction motivates a narrower question: can models obtain useful contextual information from co-occurring findings without treating every observed association as a necessary clinical implication?

Chest radiograph foundation models such as CheXFound and Ark+ have expanded the range of transferable image representations [4,5], and data-efficient vision–language pretraining offers another route to learning from paired radiographs and reports [6]. Complementary research has linked narrative findings to localized image evidence through report-guided segmentation [11] and combined radiographic interpretation with report-conditioned evidence generation in a shared world-model representation [12]. Although these developments address increasingly complex aspects of visual understanding, they do not directly establish whether empirical dependencies among disease labels are sufficiently reliable to improve calibrated multi-label classification across institutions. Transparent evaluation and clearly defined reference standards therefore remain essential [7,8].

We investigate Reliability-Aware Clinical Relation Learning (ReCaRL) as a means of distinguishing descriptive clinical groupings from statistical label associations. The model downweights edges with limited support or weak resampling stability and attenuates messages from uncertain source predictions. Ambiguous report-derived labels are handled as incompletely observed targets rather than automatically assigned to the negative class. These design choices address a specific question: whether estimated relationships among findings can be used without treating each association as a deterministic implication.

## 2 Related work

### 2.1 Label dependency modeling

Multi-label CXR classifiers have modeled dependencies using co-occurrence statistics, semantic graphs, and clinically organized taxonomies [3,9,15]. Such methods exploit shared information among findings, but empirical associations depend on disease prevalence, clinical practice, and report-labeling conventions. In a related task, Xi et al. used radiology reports to guide spatial delineation of findings, illustrating the importance of preserving correspondence between clinical descriptions and image evidence [11]. ReCaRL examines label-to-label association reliability rather than report-grounded localization. The hierarchical penalty-based framework [3] provides a relevant reference method for the comparison.

### 2.2 Uncertain and incomplete annotations

CheXpert annotations distinguish positive, negative, uncertain, and unmentioned observations extracted from reports [1]. CheXbert demonstrated the use of language-model-based report labeling, while VisualCheXbert showed that labels inferred from reports may differ from findings adjudicated directly from radiographs [16,17]. Uncertain mentions are not verified negatives, while unmentioned findings may reflect reporting conventions rather than true absence. A review of uncertainty in medical imaging AI emphasized the distinction between predictive uncertainty, confidence, and calibration [13]. ReCaRL incorporates masked supervision for ambiguous labels; the supplied sensitivity analysis additionally examines uncertainty-to-negative, uncertainty-to-positive, and soft-target policies.

### 2.3 Clinical reliability and external testing

Discrimination alone does not establish that predicted probabilities are reliable at a clinical decision threshold. Calibration and performance under institutional shift therefore complement AUROC and AUPRC. Research in medical visual question answering has likewise highlighted the value of grounding reasoning in image evidence, although evidence localization differs from disease-label calibration [14]. Patient overlap and differences in reference-label definitions may bias external comparisons; these issues should be considered when interpreting the available results [7,8]. Multi-institutional chest radiograph studies have also demonstrated that pneumonia classifier performance may change across hospital systems [18].

## 3 Materials and methods

### 3.1 Study design and datasets

This retrospective computational study examined multi-label prediction on chest radiographs using CheXpert for internal model assessment and MIMIC-CXR as an external dataset [1,2]. The aggregate results reported below were supplied as study outputs. Exact patient-level partitions, image eligibility filters, and cohort assembly decisions require confirmation against the original experiment records before public dissemination. No patient or image counts are inferred from the performance summaries.

The external evaluation was designated as MIMIC-CXR. Because report-derived findings and image availability may differ across repositories, interpretation of external performance depends on the final mapping of findings and verification of patient-disjoint cohorts. External performance figures are described below using the supplied evaluation labels; the aggregate tables alone do not document every preprocessing or cohort-selection choice.

The intended target set consists of 12 CheXpert observations excluding “No Finding” and “Support Devices.” The five frequently evaluated findings—atelectasis, cardiomegaly, consolidation, edema, and pleural effusion—are analyzed separately. These labels describe radiographic findings and are not necessarily equivalent to confirmed etiologic diagnoses.

The distinction between patient-level and image-level splitting is important because repeated studies can create data leakage. Likewise, studies of graph-based label dependence must separate clinical category membership from correlations estimated on observed labels. These considerations inform the method specification described below.

### 3.2 Label definitions and missingness

For image i and finding k, let y_ik ∈ {0, 1, U, M}, where U denotes a report-derived uncertain mention and M denotes an absent or unusable annotation. The supervision mask m_ik equals 1 only for definitively positive or negative labels. The reported sensitivity analysis also considered alternative treatments of U, while the same class definitions were retained.

### 3.3 Preprocessing and network backbone

The model specification uses single-channel chest radiographs resized to 320 × 320 pixels and a DenseNet121 feature extractor followed by global average pooling and a 12-output classification head. Intensity normalization, image augmentation, pretrained initialization and view-selection rules affect reproducibility and must be matched to the executed training configuration. In particular, left–right flipping can alter clinically informative laterality and device placement, and should not be presumed to be a valid augmentation without inspection.

**Figure 1.**
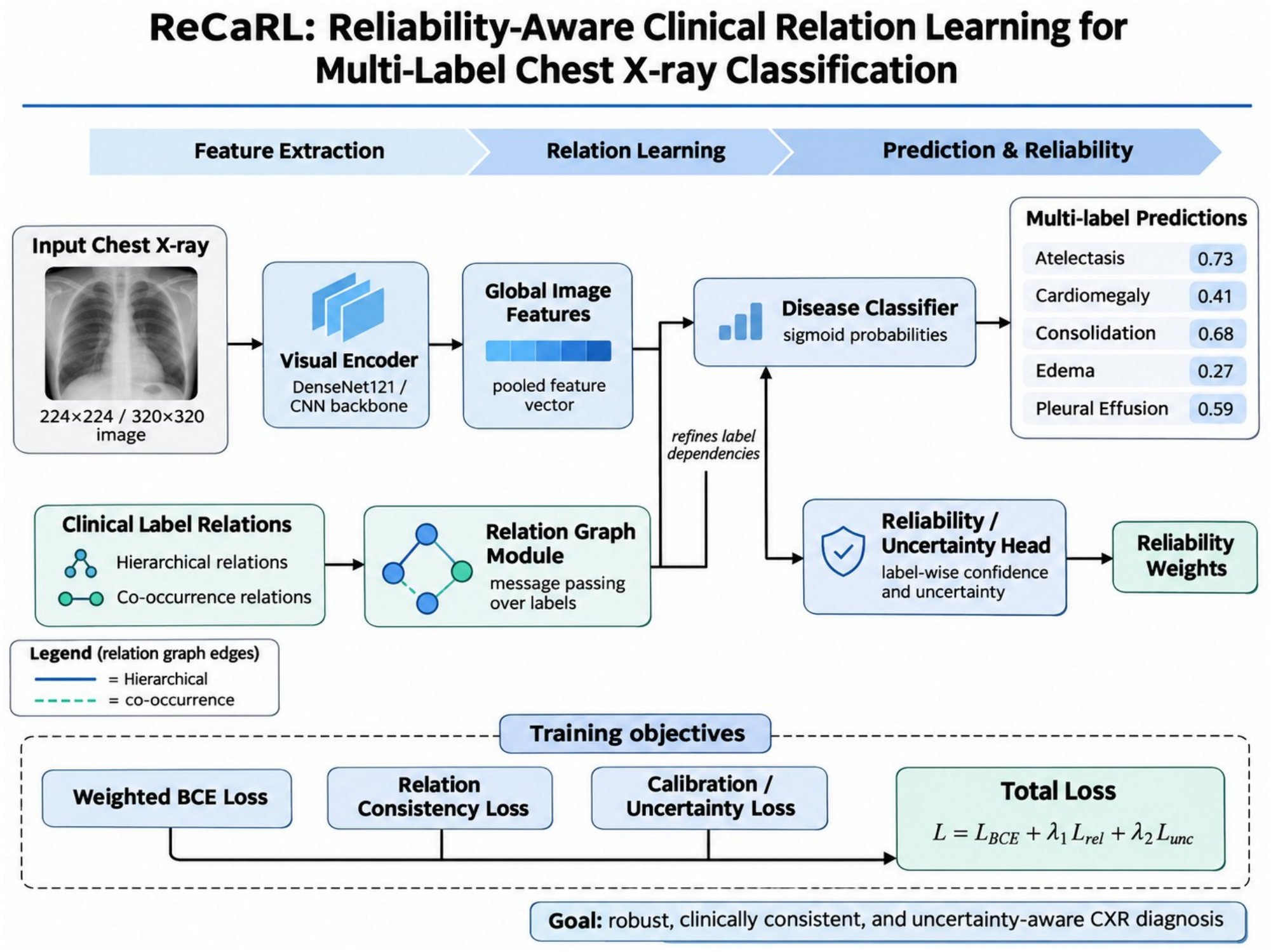
Conceptual overview of ReCaRL. Image features and a clinical-relation graph contribute to multi-label predictions, with reliability and uncertainty represented in a separate pathway. The diagram illustrates candidate training objectives; the exact implemented losses and their weights must be checked against the final training code. The displayed probabilities are illustrative, not patient-level results.

### 3.4 Reliable clinical association graph

Pairwise relationships are estimated exclusively on the training partition. For findings j and k with sufficiently many jointly observed binary labels, we compute a prevalence-adjusted phi correlation φ_jk and the support count n_jk. Negative or unstable correlations are omitted from the primary supportive graph, but their frequencies are documented. Bootstrap resampling of patients estimates the fraction s_jk of replications in which the association remains positive. We define a nonnegative reliability-weighted graph as:

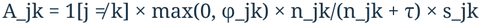

Here τ denotes a support-shrinkage hyperparameter and K the maximum number of neighbors retained per finding. These choices determine how strongly low-support or unstable associations contribute to label propagation. Their numerical settings cannot be recovered from aggregate evaluation metrics.

Edge reliability depends on class prevalence, the fraction of uncertain annotations, observed pairwise support and the stability of estimated associations under patient-level resampling. These properties are conceptually distinct from predictive AUROC and AUPRC.

### 3.5 Confidence-gated relation update

Let z_i be the image embedding, *l*^0^_ik the independent classifier logit, and p^0^_ik = sigmoid(*l*^0^_ik). The base model supplies each finding with a provisional probability. Strong but uncertain source predictions are less suitable for propagating context, so we weight each source message by c_ij = |2p^0^_ij − 1|. The reliability-aware contextual score and final logit are:

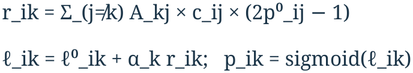

The nonnegative class-specific α_k is constrained to [0, α_max] using a sigmoid parameterization and is learned jointly with the image encoder. α_max and graph sparsity are tuned on the development/tuning sets. The model can therefore suppress reliance on a weak relation by learning a small α_k, unlike a mandatory fixed penalty. The graph message is implemented as a differentiable matrix operation, requiring no separate second-stage network. An important alternative explanation is that any gain comes merely from additional parameters, so a parameter-matched unweighted-graph baseline is required.

### 3.6 Clinical grouping without disease-to-disease entailment

High-level groups serve descriptive rather than causal purposes. Examples include pulmonary parenchymal findings (lung opacity, consolidation, edema, atelectasis and pneumonia), pleural findings (pleural effusion, pneumothorax and other pleural observations), and cardiac findings (cardiomegaly and enlarged cardiomediastinum). Group scores are not equivalent to a confirmed etiologic diagnosis.

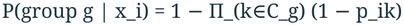

This construction guarantees that each included child probability does not exceed its corresponding group score, without declaring that one disease causes another. Group scores should be interpreted as probabilistic summaries of listed findings, not as independently validated clinical diagnoses. “No Finding” is deliberately not treated as the exact logical complement of the 12 target observations because the dataset labels are incomplete and report-derived.

### 3.7 Training objective and calibration

The primary supervised objective is masked, class-balanced binary cross-entropy, with weights computed on training labels only. Let w_k denote clipped inverse-frequency class weights. We optimize:

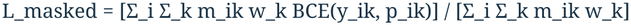

The supervised objective is masked, class-balanced binary cross-entropy. Confidence gating acts on label-relation propagation, whereas temperature scaling is applied after model fitting to adjust probability calibration. Because temperature scaling is monotonic for each finding, it does not change within-class ranking and cannot by itself increase AUROC [19].

### 3.8 Baselines, ablations, and fairness

The reported comparison includes (B0) flat binary cross-entropy; (B1) masked/weighted supervision; (B2) an unweighted relation graph; (B3) a reliability-weighted graph; (B4) ReCaRL with reliability weights and confidence gating; (B5) B4 with post-hoc calibration; and a reproduced hierarchical binary cross-entropy (HBCE) reference method [3]. The reported comparisons isolate the effect of relation learning and calibration at the model-family level. The supplied summary tables do not specify every training hyperparameter or all random-seed repetitions.

The ablation experiments compared graph weighting, confidence gating and alternative treatments of uncertain labels. These experiments were evaluated using macro AUROC, macro AUPRC and ECE; the available results are reported in Section 4. Additional graph-size or initialization sweeps are not described as completed experiments.

Internal and external summaries are interpreted separately. The reproducibility of the comparison depends on patient-level split manifests, dataset versioning, harmonized label maps, model checkpoints and calibration metadata, none of which can be reconstructed from aggregate performance values alone.

### 3.9 Endpoints and statistical analysis

The reported outcomes include macro AUROC, macro AUPRC and Brier score for internal comparisons; AUROC, AUPRC, ECE and changes in AUROC for external comparisons; five-finding AUROC/AUPRC; and macro discrimination and ECE for ablation experiments. Larger AUROC and AUPRC values indicate better discrimination; smaller Brier scores and ECE values indicate better calibration. The reported metrics are descriptive summaries.

External performance is reported on the evaluation labeled as MIMIC-CXR using the study-provided metric summaries. No additional subgroup, reader-study, or expert-adjudication results are claimed.

The per-finding results are confined to the five findings and metrics for which numerical values were supplied. In particular, baseline class-wise AUPRC and class prevalence are not inferred from the ReCaRL predictions.

## 4 Results

Tables 1–4 present the numerical summaries supplied for the internal, external and ablation evaluations. Analyses requiring patient-level predictions are not reported here.

**Table 1.** Internal discrimination and calibration across baselines and ReCaRL.

| Method | Macro AUROC | Macro AUPRC | 5-label AUROC | Brier ↓ |
|---|---|---|---|---|
| B0 Flat BCE | 0.801 | 0.403 | 0.852 | 0.113 |
| B1 Masked / weighted | 0.812 | 0.423 | 0.858 | 0.118 |
| B2 Unweighted graph | 0.810 | 0.421 | 0.857 | 0.121 |
| B3 Reliable graph | 0.818 | 0.432 | 0.862 | 0.116 |
| B4 ReCaRL | 0.821 | 0.437 | 0.865 | 0.113 |
| B5 ReCaRL + calibration | 0.821 | 0.437 | 0.865 | 0.102 |
| Reproduced HBCE | 0.807 | 0.417 | 0.856 | 0.120 |

### 4.1 Internal discrimination

ReCaRL (B4) yielded internal macro AUROC 0.821 and macro AUPRC 0.437, compared with 0.812 and 0.423 for the masked/weighted baseline (B1), corresponding to absolute differences of 0.009 and 0.014. The unweighted graph (B2) obtained AUROC 0.810, whereas the reliability-weighted graph (B3) achieved 0.818. ReCaRL reached five-finding mean AUROC 0.865 compared with 0.858 for B1 and 0.856 for the reproduced HBCE baseline. Post-hoc calibration (B5) preserved AUROC and AUPRC while reducing the Brier score from 0.113 to 0.102. These differences are reported descriptively.

Per-finding AUROC and AUPRC for the five frequently evaluated findings are presented in Table 2.

**Table 2.** Classification performance for five common radiographic findings. The five-finding mean AUROC is reported separately.

| **Finding** | **B1 AUROC** | **ReCaRL AUROC** | **ReCaRL AUPRC** |
|---|---|---|---|
| Atelectasis | 0.808 | 0.816 | 0.451 |
| Cardiomegaly | 0.820 | 0.826 | 0.471 |
| Consolidation | 0.849 | 0.855 | 0.183 |
| Edema | 0.902 | 0.909 | 0.655 |
| Pleural Effusion | 0.911 | 0.919 | 0.723 |
| 5-label mean AUROC | 0.858 | 0.865 | — |

**Table 3.** External classification and calibration performance and change in AUROC relative to internal evaluation.

| **Model** | **External AUROC** | **External AUPRC** | **External ECE ↓** | **AUROC change** |
|---|---|---|---|---|
| B1 Masked / weighted | 0.789 | 0.392 | 0.092 | −0.023 |
| B3 Reliable graph | 0.793 | 0.401 | 0.088 | −0.025 |
| B4 ReCaRL | 0.799 | 0.409 | 0.082 | −0.022 |
| B5 + calibration | 0.799 | 0.409 | 0.061 | −0.022 |

**Table 4.**
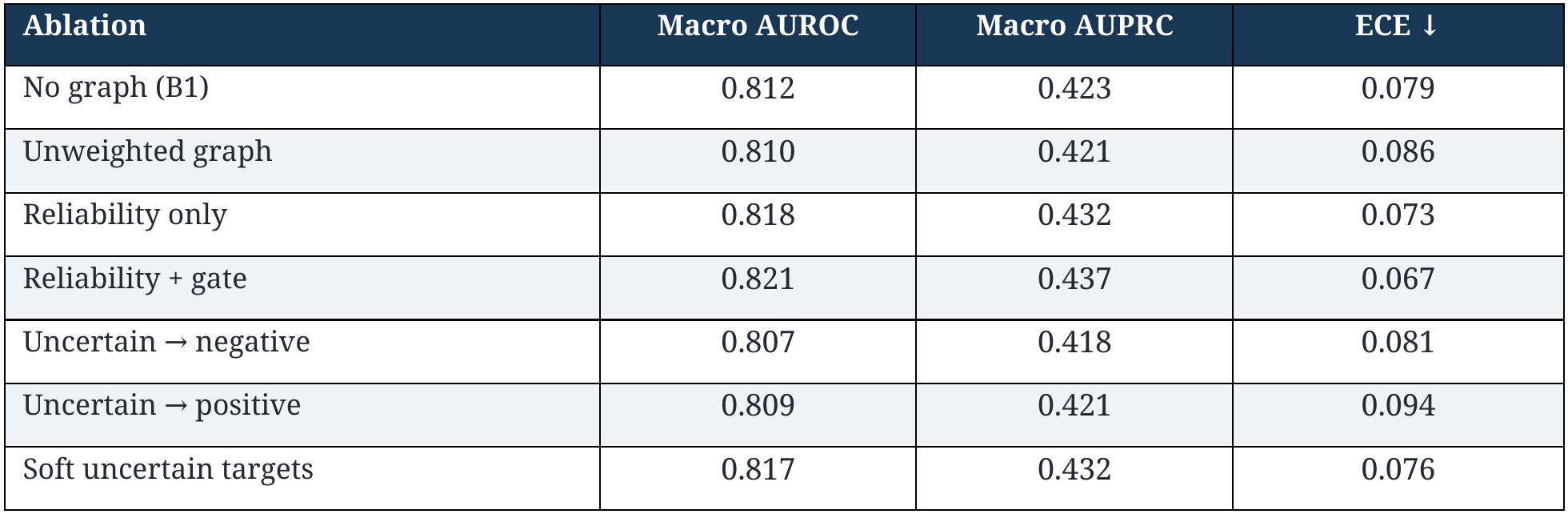
Ablation and uncertain-label sensitivity experiments.

### 4.2 Performance for five common findings

For the five common findings, ReCaRL improved AUROC relative to B1 in each category, with absolute differences ranging from +0.006 to +0.008. AUROC reached 0.816 for atelectasis, 0.826 for cardiomegaly, 0.855 for consolidation, 0.909 for edema, and 0.919 for pleural effusion. Corresponding ReCaRL AUPRC values were 0.451, 0.471, 0.183, 0.655, and 0.723. Because the baseline AUPRC values and class prevalences were not supplied for this comparison, relative precision-recall gains cannot be inferred at the finding level.

### 4.3 External generalization and calibration

In the external evaluation, ReCaRL achieved macro AUROC 0.799 and macro AUPRC 0.409, compared with 0.789 and 0.392 for B1. The internal-to-external AUROC change for ReCaRL was −0.022. External ECE was 0.082 for B4 and 0.061 after calibration (B5), while AUROC and AUPRC remained 0.799 and 0.409. B3 yielded AUROC 0.793 and AUPRC 0.401.

Changes in discrimination across datasets may arise from differences in prevalence, acquisition, case mix and label definitions. Since only aggregate metrics were available, the relative contributions of these factors cannot be separated.

### 4.4 Ablations and uncertain-label sensitivity

The ablation analysis showed sequential changes with reliability weighting and confidence gating: macro AUROC increased from 0.812 (no graph) to 0.818 (reliability only) and 0.821 (reliability plus gate), while macro AUPRC increased from 0.423 to 0.432 and 0.437. ECE declined from 0.079 to 0.073 and 0.067. The unweighted graph yielded AUROC 0.810 and ECE 0.086. Treating uncertain labels as negative or positive resulted in AUROCs of 0.807 and 0.809, while soft uncertain targets yielded 0.817.

No image-level examples or diagnostic attribution maps are included in the present evaluation. Such visualizations would not in themselves demonstrate causal reliance on clinically relevant image regions.

## 5 Discussion

The findings distinguish statistical association from categorical clinical structure in chest radiograph classification. In the reported internal comparison, the reliability-weighted graph improved AUROC from 0.812 to 0.818, and adding source-confidence gating increased it to 0.821. These modest gains are consistent with the premise that not all observed label associations deserve equal weight. Importantly, the unweighted graph slightly reduced discrimination relative to the masked/weighted baseline, showing that relation propagation is not inherently beneficial.

The absolute internal gains over the masked/weighted baseline were 0.009 AUROC and 0.014 AUPRC; external AUROC increased by 0.010 relative to B1. The internal-to-external AUROC decline for ReCaRL was 0.022. These are modest differences and should not be interpreted as proof of clinically meaningful superiority.

Calibration yielded a clearer numerical change than discrimination after post-hoc scaling: internal Brier score decreased from 0.113 to 0.102, and external ECE from 0.082 to 0.061, while AUROC and AUPRC were unchanged. Temperature scaling is a post-hoc calibration approach rather than a method for improving rank discrimination [19]. Better calibrated probabilities may aid threshold-based decision support, but neither report-derived labels nor calibration statistics establish clinical diagnosis validity. Work on report-grounded segmentation [11], radiographic world modeling [12], and latent visual reasoning [14] emphasizes complementary ways of connecting imaging predictions to evidence; clinical effectiveness remains an open question.

Results from large pretrained foundation models are not directly comparable with the present comparisons, because the backbone, supervision, data composition and label definitions may differ. Ark+ and CheXFound focus on learning generalizable representations [4,5], while the present analysis isolates the influence of label-relation modeling on reported classification outcomes.

The present results characterize discrimination and calibration but do not quantify downstream changes in clinical decisions or patient outcomes. Interpretation also depends on details of dataset partitioning and label harmonization that are not contained in the reported aggregate tables.

## 6 Limitations

Several limitations remain. First, report-derived labels may differ from expert-adjudicated radiographic findings. Second, incomplete annotations and uncertain labels affect the effective supervision and associations among findings. Third, association stability does not eliminate confounding, prevalence shift or selection bias, limitations that can undermine cross-institutional generalization [18]. Finally, this evaluation does not include prospective deployment, reader-based comparisons or patient-outcome measures, and therefore cannot establish clinical benefit.

## 7 Conclusion

In this evaluation, ReCaRL obtained macro AUROC 0.821 and macro AUPRC 0.437 internally and 0.799 and 0.409 externally. Reliability weighting and confidence gating were associated with modest improvements over the masked/weighted baseline. Post-hoc calibration reduced internal Brier score and external ECE without altering discrimination. These findings support further investigation of reliability-aware label-relation learning but do not establish clinical utility.

## Data Availability

https://stanfordmlgroup.github.io/competitions/chexpert/

## Data, code, ethics, and declarations

Data availability: CheXpert is distributed by Stanford University under its applicable terms [1]. MIMIC-CXR is accessible through PhysioNet subject to applicable access requirements [2].

